# COVID-19 Infection, Admission and Death Amongst People with Rare Autoimmune Rheumatic Disease in England. Results from the RECORDER Project

**DOI:** 10.1101/2021.08.17.21260846

**Authors:** Megan Rutter, Peter C. Lanyon, Matthew J. Grainge, Richard Hubbard, Emily Peach, Mary Bythell, Peter Stilwell, Jeanette Aston, Sarah Stevens, Fiona A. Pearce

**Affiliations:** Department of Population and Lifespan Sciences, School of Medicine, University of Nottingham, Nottingham, UK; Department of Rheumatology, Nottingham University Hospitals NHS Trust, Nottingham, UK; National Congenital Anomaly and Rare Disease Registration Service, National Disease Registration Service, Public Health England, UK; National Institute for Health Research (NIHR) Nottingham Biomedical Research Centre, Nottingham, UK

**Keywords:** COVID-19, coronavirus, mortality, rare autoimmune rheumatic diseases, epidemiology, shielding, infection

## Abstract

**Objectives:** To calculate the rates of COVID-19 infection and COVID-19-related death among people with rare autoimmune rheumatic diseases (RAIRD) during the first wave of the COVID-19 pandemic in England compared to the general population.

**Methods:** We used Hospital Episode Statistics to identify all people alive 01 March 2020 with ICD-10 codes for RAIRD from the whole population of England. We used linked national health records (demographic, death certificate, admissions and PCR testing data) to calculate rates of COVID-19 infection and death up to 31 July 2020. Our primary definition of COVID-19-related death was mention of COVID-19 on the death certificate. General population data from Public Health England and the Office for National Statistics were used for comparison. We also describe COVID-19-related hospital admissions and all-cause deaths.

**Results:** We identified a cohort of 168,680 people with RAIRD, of whom 1874 (1.11%) had a positive COVID-19 PCR test. The age-standardised infection rate was 1.54 (95% CI 1.50-1.59) times higher than in the general population. 713 (0.42%) people with RAIRD died with COVID-19 on their death certificate and the age-sex-standardised mortality rate for COVID-19-related death was 2.41 (2.30 – 2.53) times higher than in the general population. There was no evidence of an increase in deaths from other causes in the RAIRD population.

**Conclusions:** During the first wave of COVID-19 in England, people with RAIRD had a 54% increased risk of COVID-19 infection and more than twice the risk of COVID-19-related death compared to the general population. These increases were seen despite shielding policies.

**Key Messages:**

1. People with RAIRD were at increased risk of COVID-19 infection during the first wave.
2. Compared to the general population, they had over twice the risk of COVID-19-related death.
3. These increased risks were seen despite shielding policies in place in England.

## Introduction

Our previous work has shown that people with rare autoimmune rheumatic diseases (RAIRD) were at increased risk of all-cause mortality during the first wave of the COVID-19 pandemic (March-April 2020), when compared to the general population in England(1). However, this study did not examine whether the increased mortality was due to COVID-19 infection itself, or due to indirect effects of the pandemic.

This study uses linked national health records for the whole population of England to calculate the rates of laboratory confirmed COVID-19 infection and COVID-19-related death among people with RAIRD from 01 March to 31 July 2020, the first wave of the COVID-19 pandemic, and compares these rates to that in the general population. We describe COVID-19-related hospital and ICU admission, underlying causes of death by category and COVID-19-related mortality stratified by RAIRD diagnosis.

## Methods

### Background

The Registration of Complex Rare Diseases Exemplars in Rheumatology (RECORDER) project is a collaboration between the University of Nottingham, Nottingham University Hospitals NHS Trust and the National Congenital Anomaly and Rare Disease Registration Service (NCARDRS). NCARDRS, based within Public Health England (PHE), registers people with rare conditions in order to support high quality clinical practice and research, provide epidemiology data and empower patients(2). It has unique access to linked national datasets of electronic health records at patient-identifiable level for the whole population of England.

This study uses Hospital Episode Statistics (HES, which contains every episode of admitted patient care in NHS hospitals in England), COVID-19 polymerase chain reaction (PCR) test results and Office for National Statistics (ONS) death certificate data.

### Data validation

Our previous work validating HES ICD-10 codes for RAIRD has shown high accuracy, with a positive predictive value was 84.7%(1). Prevalence estimates based on our findings for ANCA-associated vasculitis, systemic lupus erythematosus and scleroderma, are similar to reported population estimates(3-5).

### Study cohort

People with a diagnostic code for RAIRD in HES from 2003 onwards, resident in England, and alive 01 March 2020 were included in the study, using the same cohort as our preliminary all-cause mortality study(1). A data flow diagram is shown in Supplementary Figure 1. Vital status data from the NHS Personal Demographics Service were used to confirm whether people were alive, or to confirm date of death(6).

### RAIRD diagnoses

Participants were grouped by RAIRD diagnosis, based on their most recent diagnostic code. Where the most recent code was non-specific, for example “Renal tubulo-interstitial disorder in systemic connective tissue disorder”, earlier, more specific diagnostic codes were used where available, following the algorithm in Appendix 1.

### Death certificate data

Death certificate and underlying cause of death data (free text and ICD-10 coded) provided by the ONS were utilised. Using internationally agreed rules, ONS assigns underlying cause of death, usually based on the lowest completed line of Part 1 of a death certificate(7). Our data were examined for ICD-10 codes specific to COVID-19 (U07.1, U07.2). The free text was manually checked for keywords (“*cov”*, “*virus”* or “*19”*) which confirmed that no deaths related to COVID-19 (including misspellings) had been omitted. Whilst COVID-19-specific ICD-10 codes were not introduced until 25 March 2020(8), all deaths with a free text mention of COVID-19 occurring before that date had been captured retrospectively by the ONS coding system. We classified underlying all-cause death by category.

### COVID-19 status

A population-level dataset of COVID-19 PCR test results, held in the Second Generation Surveillance System in PHE, was used. Positive tests amongst the RAIRD cohort were extracted, along with the date the laboratory reported the result. Demographics are described by pillar 1 (in-hospital) or pillar 2 (community) testing.

### Hospital and intensive care unit admissions

HES admitted patient care (APC) data on hospital and intensive care unit (ICU) admissions with an ICD-10 diagnostic code for COVID-19 were extracted. Duration and number of admissions, and basic and advanced respiratory support days on ICU are described.

### Mortality rate calculation

We report two measures of COVID-19-related deaths. Our primary definition is death with any mention of COVID-19 on the death certificate as used by the ONS(9). This was chosen due to the limited availability of COVID-19 PCR testing in the community during the first wave. Our secondary definition is death within 28 days of a positive COVID-19 PCR test, as used by PHE(10).

The crude all-cause mortality rate from 01 March to 31 July 2020 was calculated, with the cohort of patients identified as having RAIRD used as the denominator population, along with the crude mortality rates for the two measures of COVID-19-related death.

Age-sex-standardised mortality rates per 100,000 in the population were calculated, standardised to the 2019 mid-year estimate for the England population using 5-year age bands. Age-standardised mortality rates (ASMRs) standardised to the 2013 European Standard Population (ESP) were also calculated. As the ESP is not disaggregated by sex and assumes equal numbers of males and females, and identical age distributions within sexes, this population was not used to calculate age-sex standardised rates.

The ONS provided data for all-cause deaths, and deaths with any mention of COVID-19 on the death certificate, over the same time period in England, split by sex and age band (11, 12). PHE provided comparable data for deaths within 28 days of a positive COVID-19 test (available to the public on request). These data were used to calculate the crude, age-standardised and age-specific mortality rates as a comparator. Publicly available data from the government Coronavirus dashboard(13) were used to compare infection rates in the general population. The 2019 mid-year estimate for the population of England was used as the denominator.

### COVID-19 infection rate calculation

Laboratory confirmed COVID-19 infection rate from 01 March to 31 July 2020 was calculated, with the cohort of patients identified as having RAIRD used as the denominator population. Infection rate was age-standardised to the mid-year 2019 England population.

### Stratification by disease

Poisson regression methods were used to analyse rates of COVID-19-related death, adjusted for age, sex and RAIRD diagnosis. Incidence rate ratios for each diagnosis, with 95% confidence intervals, were displayed as a forest plot.

### Ethics

This study received a favourable opinion from the Camden and Kings Cross Research Ethics Committee, study reference 20/HRA/2076, on 18 June 2020.

The legal basis to access the data is predominantly covered by NCARDRS’ Section 251 approval (Reference CAG 10-02(d)/2015). Where the work extends beyond Section 251 approval, it has been approved under Regulation 3(4) of the Health Service Control of Patient Information Regulations 2002 (COPI), allowing the processing of confidential patient information for the purposes of protecting public health and managing the COVID-19 outbreak.

For quality assurance the data extraction and analysis were re-conducted by an independent analyst from the National Disease Registration Service.

### Patient and public involvement

This work has been developed with input from people with RAIRD. Following our findings of increased mortality in this population(1), we consulted with patients and patient charities to confirm priorities for future research and inform the communication and dissemination of our results. This will continue as an iterative process as results become available. A plain english summary of this study is available as an online supplement.

### Data analysis

Cleaning, linkage and analysis of the data was performed in R version 3.6.3 (packages *tidyverse(14), janitor(15), survival(16), mfx(17), survminer(18), meta(19)*).

## Results

### Demographic data

168,691 people were included in the RAIRD cohort in our all-cause mortality study(1). Updates to personal demographic data revealed that 11 people had died prior to 01 March 2020, leaving 168,680 people included in this study. Descriptive demographic data, including RAIRD diagnoses, are shown in Table 1. The median age of the population was 61.7 years (IQR 41.5 – 75.4) and 118,374 (70.2%) were female.

**Table 1:**
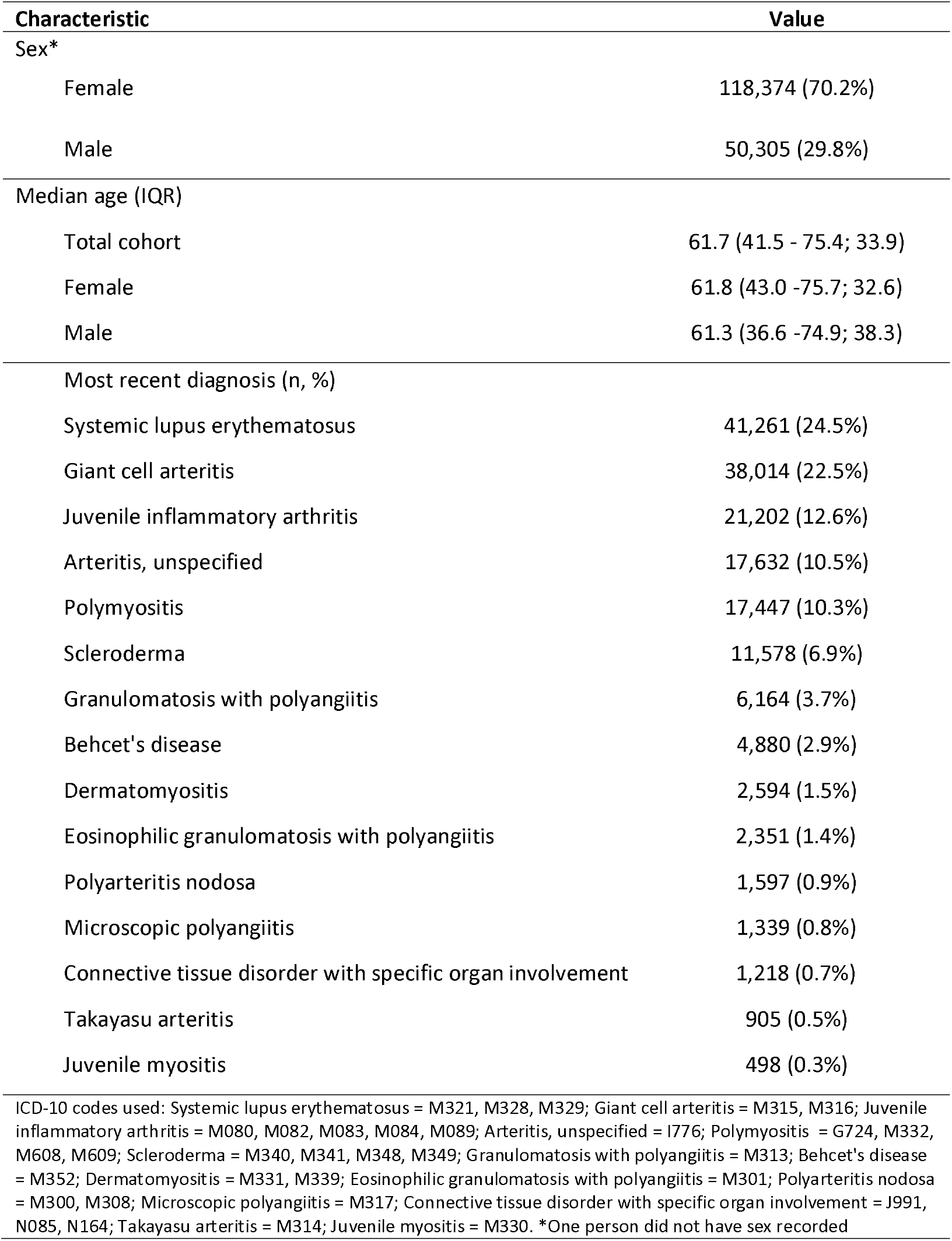
Characteristics of the cohort of people with RAIRD alive 01 March 2020 (n=168,680)

### COVID-19 infection

Between 01 March and 31 July 2020, 1874 (1.11%) of the RAIRD population had a positive COVID-19 PCR test, compared to 261,348 (0.46%) of the general population. Age-standardised to the England population, the infection rate per 100,000 person-years was 1,720.3 (1670.0-1770.6), compared to 1114.3 (1111.5-1117.0) per 100,000 person-years in the general population (rate ratio 1.54 (1.50-1.59), Table 2).

**Table 2:** PCR-proven COVID-19 infections in the RAIRD population compared to the whole population of England

|  | RAIRD (n = 168,680) | England (n = 56,286,961) | Rate ratio |
| --- | --- | --- | --- |
| Infection rate (n, %) | 1,874 (1.11%) | 261,348 (0.46%) |  |
| Death certificate mention of COVID (n, %) | 713 (0.42%) | 56,196 (0.10%) |  |
| Death within 28 days of COVID test (n, %) | 574 (0.34%) | 36,658 (0.07%) |  |
| Age-standardised COVID-19 infection rate (per 100,000 person-years) | 1,720.3 (1670.0-1770.6) | 1,114.3 (1111.5-1117.0) | 1.54 (1.50-1.59) |
| Positive COVID-19 PCR tests by pillar (n (% total tests)) | RAIRD (n = 168,680) | England (n = 56,286,961) |  |
| Pillar 1 | 1,628 (86.87%) | 164,600 (62.31%) |  |
| Pillar 2 | 246 (13.13%) | 99,583 (37.69%) |  |
| Age of those in RAIRD cohort with a positive PCR test |  |  |  |
|  | Mean age | Median age | Interquartile range |
| Pillar 1 | 71.14 | 75.10 | 60.89 – 84.14 (23.25) |
| Pillar 2 | 62.42 | 65.31 | 42.19 – 84.72 (42.53) |
| Deaths in RAIRD cohort within 28 days of a positive PCR test, by pillar (n (% deaths)) |  |  |  |
| Pillar 1 | 560 (97.6%) |  |  |
| Pillar 2 | 14 (2.4%) |  |  |
| Age of RAIRD cohort who died within 28 days of a positive PCR test |  |  |  |
| Pillar 1 | 77.03 | 79.21 | 71.26 - 85.94 (14.68) |
| Pillar 2 | 88.13 | 89.47 | 84.84 - 92.55 (7.71) |
| Note: Pillar 1 denotes in-hospital testing and Pillar 2 denotes community testing |  |  |  |

Characteristics of those with a positive COVID-19 PCR test are shown in Table 2. People with RAIRD with a positive PCR test were more likely to have had a pillar 1 test (86.87% of positive tests) than people with a positive PCR test in the general population (62.31% of positive tests).

### All-cause mortality

Between 01 March and 31 July, 3401 (2.02%) people in the RAIRD cohort died of any cause, compared to 257,547 out of 56,286,961 (0.46%) among the general population.

### COVID-19 related mortality

Of the 3401 people who died, death certificate data were available for 3332 (97.97%). 713 (0.42% RAIRD cohort) had COVID-19 mentioned on their death certificate, in any position. This compares to 49,166 (0.09%) of all those who died in the general population.

Of those with a positive COVID-19 PCR test, 574/1874 (30.6%) died within 28 days of a positive test, compared to 36,658/261,348 (14.03%) of the general population. However, the RAIRD cohort were older than the general population of England.

The combined total of people with RAIRD dying with either COVID-19 mentioned on their death certificate, or within 28 days of a positive COVID-19 test, is reported in Appendix 2.

### Age-and age-sex-standardised mortality rates

The age-sex-standardised mortality rate for all-cause death in RAIRD, standardised to the 2019 mid-year population of England, was 2325.2 (2274.8 – 2375.7), compared to 1098.1 (95% CI 1095.4-1100.9) in the general population (rate ratio 2.12 (2.07 – 2.16)). For deaths mentioning COVID-19 on the death certificate, the age-sex-standardised mortality rate was 505.5 (481.5–529.4), compared to 209.6 (208.4–210.8) in the general population (rate ratio 2.41 (2.30 – 2.53)). For deaths within 28 days of a positive COVID-19 PCR test, the age-sex-standardised mortality rate in RAIRD was 422.0 (399.7–444.3), compared to 156.3 (155.3–157.3) in the general population (rate ratio 2.70 (2.56 – 2.84)). These data are summarised in Table 3. The ASMR for all-cause death in RAIRD, adjusted to the 2013 European Standard Population is available in supplementary Table 1.

**Table 3:** Deaths, crude mortality rates, age-sex-standardised mortality rates and sex-specific age-standardised mortality rates during March to July 2020 for the RAIRD cohort compared to the mid-year 2019 population of England

|  | Number of deaths | Number of people | Person-years | Crude mortality rate per 100,000 person years | RAIRD age-sex-standardised mortality rate* | England age-sex-standardised mortality rate* | Risk ratio for mortality rates |
| --- | --- | --- | --- | --- | --- | --- | --- |
| All-cause mortality |  |  |  |  |  |  |  |
| All | 3,401 | 168,680 | 70,283 | 4,839.0 (4734.0-4944.0) | **2325.2** (2274.8 – 2375.7) | **1098.1** (1095.4 – 1100.9) | **2.12** (2.07 – 2.16) |
| Female | 2179 | 118,374 | 49,323 | 4417.9 (4298.1-4537.6) | **2131.7** (2073.9 – 2189.4) | **1069.5** (1065.7-1073.3) | **1.99** (1.94 – 2.05) |
| Male | 1222 | 50,305 | 20,960 | 5830.0 (5619.0 - 6041.0) | **2518.8** (2427.7-2610.0) | **1127.5** (1123.5-1131.4) | **2.23** (2.15 – 2.31) |
| Death with any mention of COVID-19 on the death certificate |  |  |  |  |  |  |  |
| All | 713 | 168,680 | 70,283 | 1014.4 (966.4-1062.6) | **505.5** (481.5–529.4) | **209.6** (208.4–210.8) | **2.41** (2.30 – 2.53) |
| Female | 427 | 118,374 | 49,323 | 865.7 (812.7 – 918.7) | **417.4** (391.9-443.0) | **186.2** (184.6-187.8) | **2.24** (2.10 – 2.38) |
| Male | 286 | 50,305 | 20,960 | 1164.5 (1262.4 - 1466.6) | **595.5** (551.0-640.1) | **233.6** (231.8-235.4) | **2.55** (2.39 – 2.74) |
| Death within 28 days of a positive COVID-19 test |  |  |  |  |  |  |  |
| All | 574 | 168,680 | 70,283 | 816.7 (773.6-859.8) | **422.0** (399.7-444.3) | **156.3** (155.3-157.3) | **2.70** (2.56 -<br>2.84) |
| Female | 332 | 118,374 | 49,323 | 673.1 (626.3 -<br>719.9) | **327.2** (304.5-349.9) | **131.1** (129.8-132.5) | **2.50** (2.32 -<br>2.67) |
| Male | 242 | 50,305 | 20,960 | 1154.6 (1060.7<br>- 1248.5) | **519.0** (476.8-561.2) | **182.1** (180.5-183.6) | **2.85** (2.62 -<br>3.08) |
| *Sex-specific rates are age-standardised only |  |  |  |  |  |  |  |

### COVID-19 related deaths over time

All-cause deaths in 2019 and 2020, deaths with any mention of COVID-19 on the death certificate and deaths within 28-days of a positive COVID-19 PCR test were plotted over time and are shown in supplementary Figure 2.

### Admissions

Demographic data for those who were admitted to hospital and/or to ICU with a diagnostic code for COVID-19 are shown in Table 4. The median age of those with a hospital admission was 75.07 years (IQR 62.13-83.64), and for an ICU admission 60.65 years (IQR 48.71-69.79).

**Table 4:** Summary of hospital and intensive care unit (ICU) admissions, including demographics and characteristics of stay

| Demographics |  |  |  |  |
| --- | --- | --- | --- | --- |
|  | n | Mean age | Median age | IQR |
| Any admission with COVID code | 1672* | 71.43 | 75.07 | 62.13-83.64 (21.51) |
| Death certificate mention of COVID-19 | 546† | 76.68 | 79.04 | 71.15-85.66 (14.51) |
| Death within 28 days of test | 504 | 76.73 | 78.95 | 71.26-85.65 (14.39) |
| ICU admission with COVID code | 137 | 59.79 | 60.65 | 48.71-69.79 (21.08) |
| Death certificate mention of COVID-19 | 64 | 62.72 | 61.59 | 51.47-74.97 (23.51) |
| Death within 28 days of test | 59 | 64.30 | 65.51 | 53.83-75.91 (22.07) |
| COVID positive, not admitted, all | 595 | 65.30 | 70.72 | 48.83-84.71 (35.88) |
| Death from all causes | 81 | 81.91 | 84.88 | 76.05-90.15 (14.10) |
| Death certificate mention of COVID-19 | 64 | 81.92 | 84.86 | 77.81-89.69 (11.89) |
| Death within 28 days of test | 70§ | 81.46 | 84.82 | 74.70-89.84 (15.14) |
| Mention of COVID-19 on death certificate, without admission or positive PCR | 103 | 82.53 | 84.78 | 77.44-89.40 (11.96) |
| *Of whom 1279/1672 had a positive COVID-19 PCR test |  |  |  |  |
| † Of whom 502/546 had a positive COVID-19 PCR test |  |  |  |  |
| §Of whom 61/70 had mention of COVID-19 on their death certificate |  |  |  |  |
| All hospital admissions‡ (n=1672) |  |  |  |  |
|  | Median | Mean | Range |  |
| Duration of admission (days) | 8.0 | 13.8 | 0-269 |  |
| Number of admissions per individual | 2.0 | 2.3 | 1-20 |  |
| ICU admissions‡ (n=137) |  |  |  |  |
|  | Median | Mean | Range |  |
| Basic respiratory support days | 2.0 | 3.6 | 0-30 |  |
| Advanced respiratory support days | 1.0 | 7.9 | 0-93 |  |
| Duration of admission (days) | 6.0 | 12.1 | 0-101 |  |
| Number of ICU admissions per individual | 1.0 | 1.2 | 1-3 |  |
| ‡Where an individual had more than one admission, totals are summed |  |  |  |  |

For hospital admissions, the median length of stay was 8 days (mean 13.8, range 0-269) and the median number of admissions was 2 (mean 2.3, range 1-20).

For ICU admissions, the median length of stay was 6 days (mean 12.1, range 0-101) and the median number of admissions was 1 (mean 1.2, range 1-3). The median number of basic respiratory support days whilst on ICU was 2 (mean 3.6, range 0-30) and the median number of advanced respiratory support days was 1 (mean 7.9, range 0-93) (Table 4).

### All-cause death by category

Where death certificate data were available, underlying cause of death by category was extracted and are shown in supplementary Table 2. Deaths due to cardiovascular disease were recorded in 703 (21.1%), COVID-19 652 (19.6%), malignancy 581 (17.4%), respiratory 404 (12.1%), dementia 280 (8.4%), underlying RAIRD 113 (3.4%) and non-COVID-19 infection 19 (0.6%), with the remaining 580 (17.4%) ascribed to other causes.

For comparison, data were extracted on all-cause death in people with a diagnostic code for RAIRD occurring during March-July 2016-2020 and categorised by underlying cause (Figure 1). There was no evidence of an increase in death from non-COVID-19-related causes during the pandemic.

**Figure 1:**
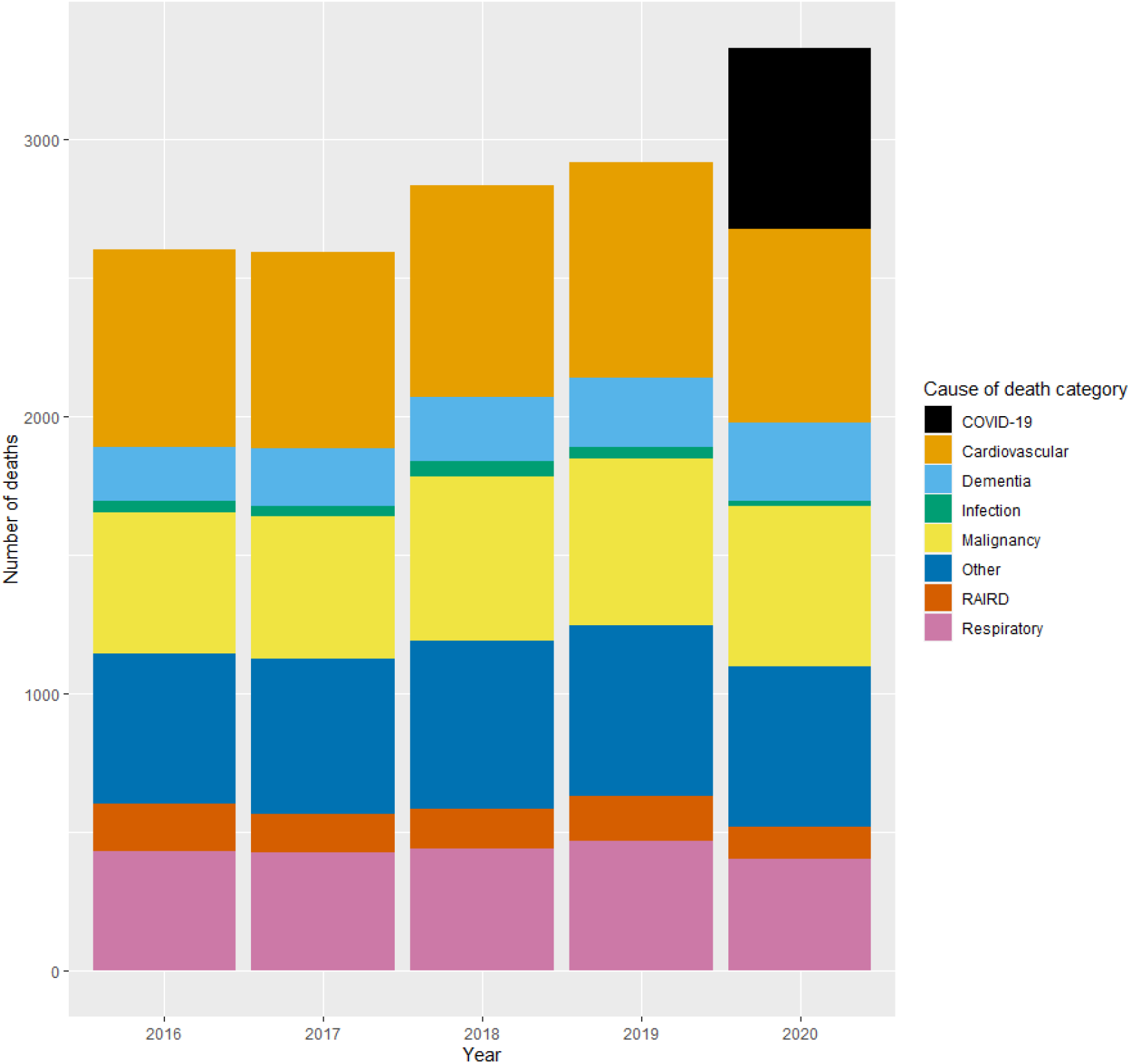
All cause death by category in people with RAIRD during March-July 2016-2020. Categories align to major ICD-10 code chapters, with the addition of diagnoses pertinent to the cohort (RAIRD, dementia).

### Age at death

There was no significant change in median age at death between 2016 and 2020 (median age ranged from 79.92-81.50), and this was similar to age at death related to COVID-19 (median 81.31, IQR 72.98-87.34; supplementary Table 3).

Age at death in people with RAIRD was compared between 2020 and 2019, categorised by underlying cause of death (supplementary Figure 3). There was no evidence of earlier age of death occurring in 2020 in certain cause of death categories e.g. in deaths from cardiovascular disease.

### Stratification by disease

Incidence rate ratios for COVID-19-related death stratified by RAIRD diagnosis are displayed in Figure 2. The comparable ratios and overlapping confidence intervals suggest a similar risk across the RAIRD cohort, regardless of diagnosis. People with giant cell arteritis (GCA) were at slightly reduced risk (RR 0.66, 95% CI 0.60-0.74), which may reflect that GCA is often a self-limiting disease not requiring lifelong immunosuppression. The wide confidence intervals reflect the relatively small number of events in the first wave.

**Figure 2:**
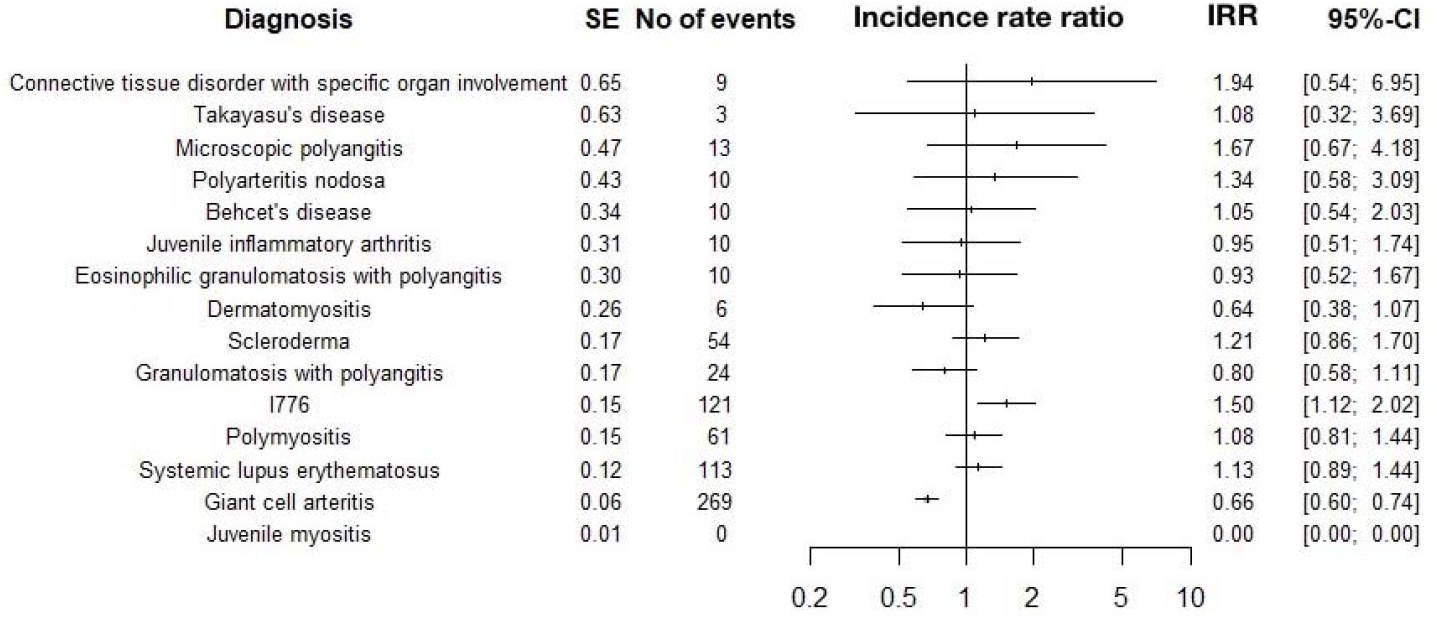
Forest plot showing incidence rate ratios with 95% confidence intervals for COVID-19-related death, stratified by RAIRD diagnosis

## Discussion

### Main findings

In the first wave of the COVID-19 pandemic, COVID-19-related death rates among people with RAIRD were more than twice that of the general population. This was contributed to by both higher COVID-19 infection rates, and higher mortality after COVID-19 infection.

The age-sex-standardised COVID-19 infection rate was 1.54 (1.50-1.59) times higher among people with RAIRD compared to the general population, despite shielding policies (protection of the clinically extremely vulnerable(20)) in place in England.

### Strengths

A major strength is that the denominator population is known, allowing us to describe for the first time both rates of COVID-19 infection and of COVID-19 related death. Previous mortality COVID-19 studies in RAIRD have either included much a smaller group of people, or relied on case series and physician reported cases(21) and allowed only internal comparisons within cohorts of people with rheumatic diseases, and not comparison with the general population.

This work uses novel linkages of HES, COVID-19 PCR testing data and death certificate data for people with rare diseases, through collaboration with NCARDRS.

Whilst our RAIRD cohort contains a heterogenous group of diseases, our sub-analysis by disease show they have comparable risks of death due to COVID-19. Pooled analysis of these diseases increases statistical power to detect outcomes and is clinically justified by their common underlying disease mechanisms and treatments.

### Limitations

The measures of COVID-19 mortality described were selected to allow comparison with available statistics for the general population. Our primary definition, any mention of COVID-19 on the death certificate (used by the ONS), infers that COVID-19 infection contributed to death, even if it was not the main cause and even in the absence of a positive PCR test. This has the theoretical potential to over-estimate deaths due to COVID-19 infection but has been found to be the best measure to explain the excess deaths seen in the national data. In our RAIRD cohort, of the 713 people with a mention of COVID-19 on the death certificate, 661 (93%) had COVID-19 in part 1, meaning that it directly lead to death(22). Conversely, we may have underestimated COVID-19-related deaths as death certificate data were unavailable for 2% of our cohort. This proportion is not unexpected and registration delays are common(23), although they may disproportionately reflect certain groups such as deaths in hospital, or due to occupational exposure to disease (including COVID-19(24)).

Our secondary definition was death within 28 days of a positive COVID-19 PCR test, as used by PHE. This measure overlooks deaths where the person never has a positive PCR test, or where the death occurred more than 28 days after diagnosis: important given the prolonged clinical course of COVID-19. In our RAIRD cohort, the median time between a positive test and death was 6 days, and only 22 (4%) of those who had COVID-19 on their death certificate, and had a positive PCR test, had that test more than 28 days prior to death.

We identified our cohort from diagnoses in HES admitted patient care (APC) data and our methodology may not have identified patients with RAIRD who have never had an in-patient or daycase admission for any reason, and who have been treated entirely on an outpatient basis. Due to the nature of RAIRD, we believe this to be the minority of cases, which is supported by disease prevalence in our cohort being similar to previous studies(3, 5, 25). However, the potential impact is to skew our cohort towards those with more severe disease.

This work describes outcomes in the first wave only. Further work is needed to describe the outcomes in the second wave, and assess the influence of immunosuppression, shielding policies and the vaccination programme.

### Comparison to the published literature

The small numbers of people affected by each RAIRD makes it difficult for studies to have enough statistical power to assess outcomes. Most research has therefore included more common rheumatic diseases such as rheumatoid arthritis (RA).

The COVID-19 Global Rheumatology Alliance describes COVID-19-related mortality in RAIRD and reports an association between immunosuppressive medications and mortality(26). However, this relied on physician-reported cases and could not report rates of COVID-19 infection or deaths.

OpenSAFELY and QCOVID studied smaller populations. At the time of the last published analysis, OpenSAFELY included 24 million patients (27) and QCOVID 8.26 million(21). Our research uses the whole England population of 56 million, which has afforded us the statistical power to calculate more precise results. The use of whole population data, with a known denominator population, also allows calculation of rates and comparison to the general population.

In addition, of the RAIRD, only systemic lupus erythematosus was included in the QCOVID and OpenSAFELY studies, in both cases combined with RA. Both found a modest increase in the risk of death (HR 1.32 (1.06-1.65) and 1.20 (1.12-1.28) respectively(27, 28)).

Whole population data from Denmark(29) on COVID-19 hospital admissions, including 10,749 people with RAIRD, supports our findings of increased risk in connective tissue disease and vasculitis (age-sex adjusted HR 1.63 (0.78–3.43) and 2.03 (1.02–4.08) respectively). The wide confidence intervals reflect the small number of events, as well as their smaller population of 4.54 million. They do not report on COVID-19 infection rates, nor on mortality in RAIRD specifically.

Whole population data from South Korea(30), including 1896 people with RAIRD, also found an increased risk of COVID-19 infection and hospitalisation (HR 1·33 (1·02–1·74) and 1·71 (1·06–2·71) respectively). There was also a suggestion of increased COVID-19 mortality but the small number of events (7 deaths) gave wide confidence intervals, which were not statistically significant (HR 1·87 (0·71–4·85)).

### Infection rate

There was an increased age-standardised infection rate in the RAIRD cohort, despite the intent of shielding policy to reduce infection exposure. There are several potential reasons for this. They may be more susceptible to symptomatic COVID-19 infection due to their underlying disease and immunosuppression. During the first wave, where testing was predominantly in hospital, their increased contact with healthcare services may have led to increased testing and ascertainment bias. People with RAIRD may have a worse state of health than the general population, leading to increased hospital admissions, necessitating COVID-19 admission screening tests and the risk of nosocomial infection. Requirements for hospital attendances may also have increased the risk of acquiring infection.

### Clinical and policy implications and future research

Our findings provide clear evidence that during the first wave of the pandemic people with RAIRD were both at higher risk of COVID-19 infection and of COVID-19-related death than people of the same age and sex in the general population. These findings have important implications for people living with RAIRD, their clinicians and for public health policy.

They confirm at whole population-level that the assumptions at the start of the pandemic, that many people with RAIRD would be clinically extremely vulnerable to the effect of COVID-19(31), were correct. Many people with RAIRD require lifelong immunosuppression, conferring ongoing risk. Protecting the health of people with RAIRD needs specific public health prioritisation, to reduce both their risk of acquiring COVID-19 infection and mortality.

Within the UK, many patients with RAIRD were classified as “clinically extremely vulnerable” to COVID-19 infection and asked to “shield”(20, 32, 33). Our findings suggest that despite this, COVID-19 infection and mortality rates were greater for people with RAIRD than for the general population. This is observational data, and the shielding status of this cohort is not yet known, so it is not possible to determine what the outcomes would have been without shielding policies in place. In addition, shielding policies were instituted on 23 March 2020 and some of our findings may reflect earlier transmission. Further research in this cohort during later waves of the pandemic, including assessment of the impact of shielding, is ongoing.

We did not find an excess of non-COVID-19-related deaths during this period. This may reflect efforts to prioritise access to urgent care. However, any detrimental effect of the pandemic may have a delayed impact on mortality.

Our results highlight the urgent need for analysis of the real-world effectiveness of vaccination among people with RAIRD, given the evidence that they may respond less well to vaccination(34). This will have crucial implications for their ongoing health protection needs, including deciding the optimal vaccination schedule to maximise protection.

## Supporting information

Plain English Summary

## Data Availability

Due to legal and ethical considerations, supporting data cannot be made openly available. However, NCARDRS data are available to all who have a legal basis to access them. Further details about the data and conditions for access are available by application to the National Disease Registration Service (https://www.gov.uk/guidance/the-national-congenital-anomaly-and-rare-disease-registration-service-ncardrs). Information on how to access this data can also be obtained from the University of Nottingham data repository: DOI: 10.17639/nott.7131

## Acknowledgements

We would like to thank Chetan Mukhtyar, Reem Al-Jayyousi, Bridget Griffiths, Richard Watts, Mithun Chakravorty, Cattleya Godsave, Julie Battista, Robin Glover, Matthew Bell and Kay Randall for their help confirming diagnoses in hospital medical notes.

We would also like to thank Charlotte Eversfield for the thorough quality assurance of the data included in this study.

This work uses data that has been provided by patients, the NHS and other health care organisations as part of patient care and support. The data is collated, maintained and quality assured by the National Congenital Anomaly and Rare Disease Registration Service, which is part of Public Health England (PHE).

