## Supplementary material for "COVID-19 Infection, Admission and Death Amongst People with Rare Autoimmune Rheumatic Disease in England. Results from the RECORDER Project": Plain English Summary

### Plain english summary: Infection rates and outcomes from COVID-19 Amongst People with Rare Autoimmune Rheumatic Disease in England. Results from the RECORDER Project.

**Background**

We are a team of doctors and researchers from the RECORDER project (Registration of Complex Rare Diseases Exemplars in Rheumatology), working with the University of Nottingham, Nottingham University Hospitals NHS Trust and the National Disease Registration Service at Public Health England. We used electronic health records that cover the whole of England for this research.

Our earlier research had shown that people with rare autoimmune rheumatic diseases such as vasculitis, lupus, scleroderma, juvenile idiopathic arthritis, myositis and Behcet’s disease were more likely to die, from any cause, during the first two months of the pandemic. However we weren’t sure why this was happening.

We looked at whether people with rare autoimmune rheumatic diseases were more likely to have COVID-19 infection and whether they were more likely to die of COVID-19 compared to people from the general population during the first wave of the COVID-19 pandemic.

**Our findings**

We studied nearly 170,000 people in England with rare autoimmune rheumatic diseases. Between March and July 2020, during the first wave of the COVID-19 pandemic in England, we found that:

- 1874 (1.11%) had COVID-19 infection (PCR test positive)
- Taking age into account, the infection rate in people with rare autoimmune rheumatic diseases was 54% higher than in the general population
- The increased infection rate occurred despite shielding policies
- 713 (0.42%) people living with rare autoimmune rheumatic disease died related to COVID-19 infection
- Taking age and sex into account, COVID-19 related death was 2.4x more common in people with rare autoimmune rheumatic disease compared to the general population
- During this time period, there was no evidence of an increase in deaths from other causes, such as heart attacks. However, it may be too soon to detect any change.

**Implications for health policy**

Our results confirm that many people with rare autoimmune rheumatic diseases are at increased risk of COVID-19 infection and COVID-19-related death. Protecting the health of people with rare autoimmune rheumatic diseases, many of whom are immunosuppressed, needs to have a higher prioritisation in public health policy. The impact of immunosuppression, and of shielding, is a focus of ongoing research.

Our results highlight the urgent need to understand the real-world effectiveness of vaccination among people with rare autoimmune rheumatic diseases, as there are concerns that they may respond less well to vaccination than the general population.
