## Supplementary data for "COVID-19 Infection, Admission and Death Amongst People with Rare Autoimmune Rheumatic Disease in England. Results from the RECORDER Project"

| Supplementary table 1: Deaths and age-standardised mortality rates during March to July 2020 for the RAIRD cohort compared to the 2013 European Standard Population | | | | | | | |
| --- | --- | --- | --- | --- | --- | --- | --- |
|  | Number of deaths | Number of people | Person-years | Crude mortality rate per 100,000 person years | RAIRD age-standardised mortality rate | General population age-standardised mortality rate | Risk ratio for mortality rates |
| All-cause mortality | | | | | | |  |
| All | 3,401 | 168,680 | 70,283 | 4,775.3 (4614.8-4935.8) | **2,454.4** (2401.1-2507.6) | **1,143.5** (1140.7-1146.4) | **2.15 (2.10 – 2.19)** |
| Death with any mention of COVID-19 on the death certificate | | | | | | |  |
| All | 713 | 168,680 | 70,283 | 1014.4 (966.4-1062.6) | **478.6** (456.0-501.3) | **218.0** (216.7-219.2) | **2.20 (2.09 – 2.30)** |
| Death within 28 days of a positive COVID-19 test | | | | | | | |
| All | 574 | 168,680 | 70,283 | 816.7 (773.6-859.8) | **393.1** (372.3-413.8) | **162.6** (161.5-163.7) | **2.42 (2.29 – 2.54)** |

| Supplementary Table 2: ONS ascribed underlying cause of death by category in RAIRD cohort between 1^st^ March and 31^st^ July 2020 | |
| --- | --- |
| Cause of death | n (%) |
| Category |  |
| Cardiovascular | 703 (21.1%) |
| COVID-19 | 652 (19.6%) |
| Malignancy | 581 (17.4%) |
| Other | 580 (17.4%) |
| Respiratory | 404 (12.1%) |
| Dementia | 280 (8.4%) |
| Underlying RAIRD | 113 (3.4%) |
| Non COVID-19 infection | 19 (0.6%) |

| Supplementary Table 3: Age at death of RAIRD cohort March-July 2016-2020 | | |
| --- | --- | --- |
| Year | Median age | IQR |
| 2020 |  |  |
| All deaths | 81.50 | 72.95-87.82 (14.87) |
| COVID-related | 81.31 | 72.98-87.34 (14.36) |
| Non-COVID-related | 81.59 | 72.92-87.92 (15.01) |
| 2019 | 81.06 | 71.72-87.07 (15.35) |
| 2018 | 79.92 | 71.15-86.99 (15.84) |
| 2017 | 80.77 | 71.05-87.20 (16.15) |
| 2016 | 80.50 | 71.67-86.94 (15.27) |

Supplementary Figure 1: Data flow diagram showing identification of RAIRD cohort from HES data.

*FCE = finished consultant episode. HES = hospital episode statistics.*

HES inpatient data interrogated for FCEs with RAIRD ICD-10 codes* in any diagnosis position in any financial year 2003 onwards

N= 1,795,048

Limited to people with an NHS number

N=1,790,932

Records with NULL NHS number removed

N=4,116

Limited to people with one NHS number

N=1,788,419

10,113 records with NULL NHS number

5,997 replaced with NHS number from other records with same HES identifier

Records with same HES identifier but multiple NHS numbers removed

N=2,513

Records combined into one record per NHS number

N=1,540,139

Limited to one record per person N=248,280

Limited to people with a valid NHS number & DOB

N=247,602

NHS numbers & DOB combinations that could not be linked by the NHS Patient Demographics Service to valid patient records removed

N=678

Limited to people alive on 1 March 2020

N=168,680

People who died before 1 March 2020 removed

N=78,922

*RAIRD ICD-10 codes include: M313, M317, M301, M314, I776, M352, M315, M316, M321, M330, M332, M331, M339, M340, M341, M348, M349, M083, M084, M082, M080, M300, M308, J991, N085, N164, M328, M329, M609, G724, M608, M089

Supplementary Figure 2: Deaths in RAIRD cohort between 01 March 2020 and 31 July 2020, shown as all deaths, deaths with any mention of COVID-19 on the death certificate and deaths within 28-days of a positive COVID-19 PCR test, with all deaths in RAIRD cohort between 01 March 2019 and 31 July 2019 as a comparator

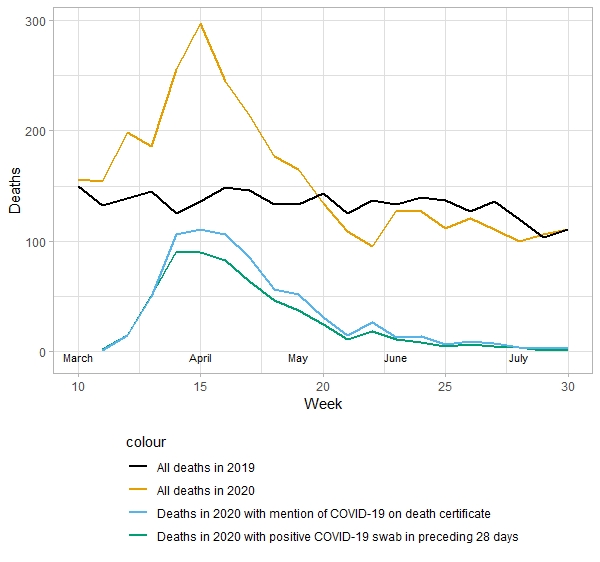

Supplementary Figure 3: Cause of death by category and age in i) between March to July 2020 and ii) mean over March to July 2016-2019

i)
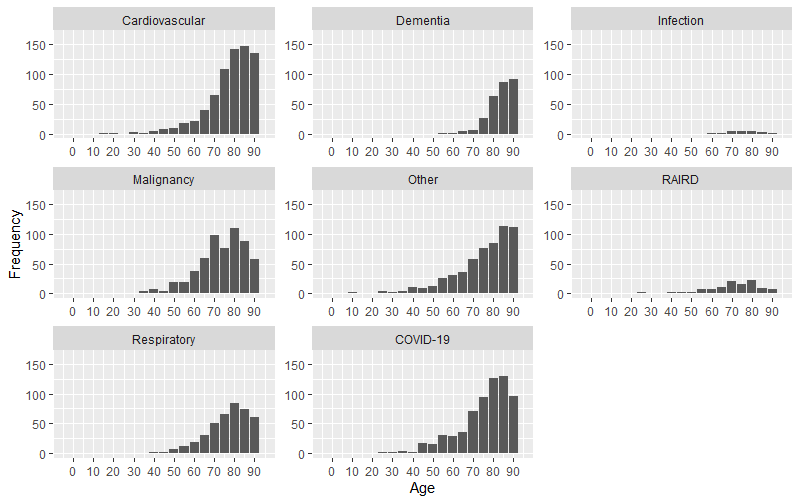

ii)

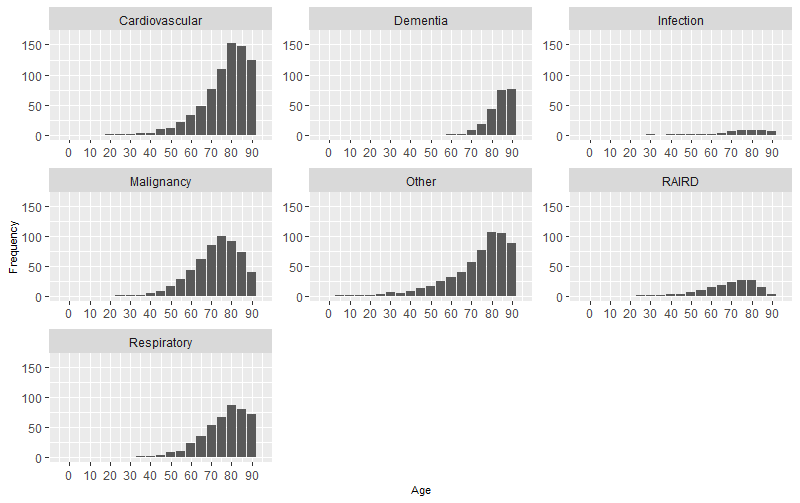

Appendix 1:

Algorithm for assigning main rheumatological diagnosis

Where primary diagnosis was a non-specific CTD code

(“Glomerular disorder in systemic connective tissue disorder”, “Renal tubulo-interstitial disorder in systemic connective tissue disorder”, “Respiratory disorder in other diffuse connective tissue disorder”)

Replaced with next most recent specific diagnostic code

Most recent diagnostic code for RAIRD applied as primary RAIRD diagnosis

Process repeated 3 times (until no further changes with repeated cycles)

Where primary diagnosis was ”Polyarteritis Nodosa” or “Arteritis, unspecified” (I776), replaced with next most recent specific diagnostic code (process not applied where next most recent code was a non-specific CTD code)

All non-specific CTD codes grouped into category “Connective tissue disorder with specific organ involvement”

Appendix 2:

The combined total of people with RAIRD dying with either COVID-19 mentioned on their death certificate, or within 28 days of a positive COVID-19 test, was 743/168,680 (0.44%) and by this measure COVID-19 was implicated in 743/3401 (21.9%) of all deaths during this time-period in this cohort. There is no similar data for the general population of England with which to compare this.
